# Multicenter evaluation of a fully automated high-throughput SARS-CoV-2 antigen immunoassay

**DOI:** 10.1101/2021.04.09.21255047

**Authors:** Dominik Nörz, Flaminia Olearo, Stojan Perisic, Matthias F. Bauer, Elena Riester, Tanja Schneider, Kathrin Schönfeld, Tina Laengin, Marc Lütgehetmann

## Abstract

**Introduction:** Molecular testing for SARS-CoV-2 continues to suffer from delays and shortages. Antigen tests have recently emerged as a viable alternative to detect patients with high viral loads, associated with elevated risk of transmission. While rapid lateral flow tests greatly improved accessibility of SARS-CoV-2 detection in critical areas, their manual nature limits scalability and suitability for large-scale testing schemes. The Elecsys^®^ SARS-CoV-2 Antigen assay allows antigen immunoassays to be carried out on fully automated high-throughput serology platforms.

**Methods:** A total of 3139 nasopharyngeal and oropharyngeal swabs were collected at 3 different testing sites in Germany. Swab samples were pre-characterized by RT-qPCR and consecutively subjected to the antigen immunoassay on either the cobas e 411 or cobas e 801 analyzers.

**Results:** Of the tested respiratory samples, 392 were PCR positive for SARS-CoV-2 RNA. Median concentration was 2.95×10^4^ (interquartile range [IQR] 5.1×10^2^–3.5×10^6^) copies/mL. Overall sensitivity and specificity of the antigen immunoassay were 60.2% (95% confidence interval [CI] 55.2–65.1) and 99.9% (95% CI 99.6–100), respectively. A 93.7% (95% CI 89.7–96.5) sensitivity was achieved at a viral RNA concentration ≥10^4^ copies/mL (∼cycle threshold (Ct) value<29.9).

**Conclusion:** The Elecsys SARS-CoV-2 Antigen assay reliably detected patient samples with viral loads of 10,000 copies/mL and higher. It thus represents a viable high-throughput alternative for screening of patients, or in situations where PCR testing is not readily available.

**Key Summary Points:** *Why carry out this study?:*

- The SARS-CoV-2 pandemic has led to a surge in demand for reliable, mass diagnostic tests worldwide.
- A thorough clinical evaluation of a fully automated high-throughput Elecsys^®^ SARS-CoV-2 Antigen assay on a total of 3139 clinical samples pre-characterized by quantitative RT-PCR was carried out.

*What was learned from the study?:*

- The assay demonstrated excellent specificity (99.9%) and good relative sensitivity, with an overall sensitivity of 60.2% and a sensitivity of 93.7% for samples containing ≥10^4^ viral RNA copies/mL.
- The Elecsys SARS-CoV-2 Antigen assay is a viable high-throughput, automated alternative to manual lateral-flow antigen tests.

## INTRODUCTION

RT-qPCR remains the gold standard for detection of SARS-CoV-2 in clinical specimens due to its unparalleled analytic accuracy [1, 2]. However, the sudden surge in demand for molecular testing continues to overstretch the capacity of both diagnostic laboratories and reagent suppliers, leading to reporting delays and sometimes inadequate availability of testing where it is urgently needed. SARS-CoV-2 antigen immunoassays have recently emerged as an alternative to nucleic acid amplification tests, most prolifically in the form of rapid, point-of-care, lateral flow tests (LFTs) [3-5]. Despite their inherent disadvantage in sensitivity compared with PCR, mathematical modeling suggests that tests could be effective for infection control if appropriate testing frequency were adopted [6]. Using currently available rapid antigen tests for mass testing does however bring about additional challenges, as this assay format is highly manual in nature and largely unsuitable for automation. Moving the SARS-CoV-2 antigen assay from LFT to a high-throughput immunoanalyzer using electrochemiluminescence detection technology could improve those issues, while further increasing overall testing capacity. The aim of this study was to evaluate the sensitivity and specificity of the new fully automated Elecsys^®^ SARS-CoV-2 Antigen assay, using samples characterized by the gold standard: RT-qPCR.

## METHODS

### Samples

In this multicenter study, nasopharyngeal and oropharyngeal samples were tested following routine diagnostics at the University Medical Center Hamburg-Eppendorf, the Hospital of Stuttgart, and Hospital Ludwigshafen (**Table S1**). Overall, the study comprised a total of 3139 nasopharyngeal and oropharyngeal swab samples, including 1331 samples from the Hospital of Stuttgart, 1058 samples from University Medical Center Hamburg-Eppendorf, and 647 samples from Hospital Ludwigshafen, as well as 103 banked nasopharyngeal samples in universal viral transport (UVT) system from a commercial vendor (Boca Biolistics, Pompano Beach, FL, USA). Specimens were collected in November 2020, in 3 mL of Copan Universal Transport Medium (UTM-RT, Copan, Brescia, Italy) or BD Universal Viral Transport (UVT, Becton, Dickinson, Sparks, MD, USA) using flocked swabs.

This study was conducted in accordance with applicable regulations, the study protocol provided by Roche Diagnostics, and the principles of the Declaration of Helsinki. The use of anonymized remnant samples material was approved by ethical review committees prior to study initiation (ethics committee names and approval numbers: Ethik-Kommission bei der Landesärztekammer Baden-Württemberg; F-2020-154 [Stuttgart]; Ethik-Kommission der Ärztekammer Hamburg: WF-184/20 [Hamburg]; Ethik-Kommission bei der Landesärztekammer Rheinland-Pfalz: 2020-15449[Ludwigshafen]).

### Antigen Assay and RT-qPCR

Antigen detection was performed at two testing sites (University Medical Center Hamburg-Eppendorf (UKE) and testing site of Augsburg) using the Elecsys SARS-CoV-2 Antigen assay on the cobas e 411 and cobas e 801 immunoanalyzers. The Elecsys SARS-CoV-2 Antigen assay (Roche Diagnostics International Ltd, Rotkreuz, Switzerland) is an automated electrochemiluminescence immunoassay (ECLIA), developed for *in vitro* qualitative detection of SARS-CoV-2 antigen. It utilizes monoclonal antibodies directed against the SARS-CoV-2 nucleocapsid protein in an antibody sandwich assay format for the detection of SARS-CoV-2 in upper respiratory tract specimens [7]. The time to result for the assay can be as little as 18 minutes, and the analyzer automatically calculates a cut-off based on the measurement of two calibrators: one negative (COV2AG Cal1) and one positive (COV2AG Cal2). The results obtained are interpreted as recommended by the manufacturer as either ‘reactive’ or ‘non-reactive’ in the form of a cut-off index (COI), i.e. ‘non-reactive’ if COI <1.0 or ‘reactive’ if COI ≥1.0.

RT-qPCR testing was performed with the qualitative cobas^®^ SARS-CoV-2 assay on the cobas^®^ 6800 system (Roche Molecular Systems, Inc., Branchburg, NJ, USA), according to manufacturer’s instructions [8]. Quantification was performed using cycle threshold (Ct) values of Target-2 (envelope protein coding gene) [9] using reference material by Qnostics Ltd. (Glasgow, UK) as the quantification standard. The formula for conversion Ct value to log10 copies/mL is (Ct × -0.30769) +13.2. The lower limit of quantification was set to Ct=33 corresponding to 1000 copies SARS-CoV-2 RNA/mL, in accordance with previous studies [9]. It has to be noted that the PCR test and quantification standard used for generating quantitative results were not recommended for this purpose by the respective manufacturers and reliability is expected to be lower than commercial quantitative solutions once they become available, which represents a limitation of this study.

### Statistical Analysis

Sensitivity and specificity, including 95% confidence intervals (CIs), were assessed as per Altman [10]. Pearson correlation was used for linear regression. The significance threshold was set at a two-sided alpha value of 0.05. Statistical analysis was performed with STATA (version 15) and GraphPad Prism (version 86 9.0.0).

### Data availability

Qualified researchers may request access to individual patient level data through the clinical study data request platform (https://vivli.org/). Further details on Roche’s criteria for eligible studies are available here: https://vivli.org/members/ourmembers/. For further details on Roche’s Global Policy on the Sharing of Clinical Information and how to request access to related clinical study documents, see here: https://www.roche.com/research_and_development/who_we_are_how_we_work/clinical_trials/our_commitment_to_data_sharing.htm.

## RESULTS

A total of 3139 respiratory samples were analyzed in this study (**Table S1**). Three hundred and ninety-two samples were SARS-CoV-2 RNA positive and 2747 samples were SARS-CoV-2 RNA negative. For PCR-positive samples, 79/392 (20.2%) patients were asymptomatic and 79.8% (313/392) were symptomatic, with 47.6% (149/313) of samples collected within the first 5 days of symptom onset. The median concentration of viral RNA was 2.95×10^4^ SARS-CoV-2 copies/mL (interquartile range [IQR] 5.1×10^2^–3.5×10^6^ SARS-CoV-2 RNA copies/mL; **Fig. 1a**). In total, 236/392 samples were detected as reactive for SARS-CoV-2 antigen with the Elecsys assay, with a median COI value of 46.0 (IQR 6.0–701.3). Distribution of the results for the PCR-positive and PCR-negative groups is shown in **Fig. 1b**. The overall sensitivity and specificity were 60.2% (95% CI 55.2–65.1, number of samples [N]=392) and 99.9% (95% CI 99.6–100, N=2747), respectively (**Table 1**). The cumulative sensitivity was 93.7%, 100%, and 100% for samples with >10,000 copies (∼Ct <29.9), 100,000 copies (∼Ct <26.6), and 1,000,000 copies (∼Ct <23.0) (**Table 1**), respectively.

**Figure 1.**
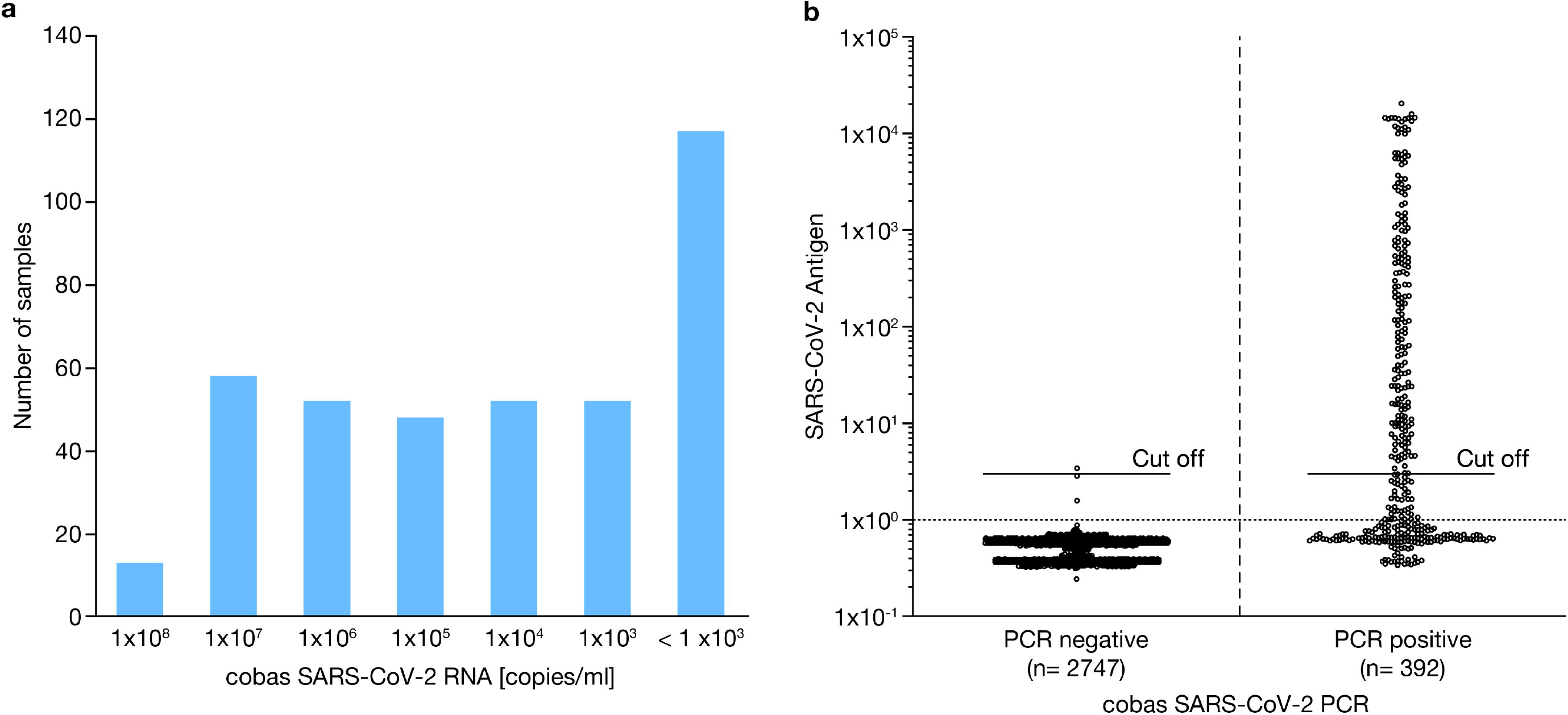
**a)** Number of positive samples relative to cobas SARS-CoV-2 PCR titer [copies/mL]; **b)** COI values of Elecsys SARS-CoV-2 Antigen test from 2747 cobas SARS-CoV-2 PCR-negative and 392 cobas SARS-CoV-2 PCR-positive samples.

**Table 1.** Sensitivity and specificity results for the Elecsys SARS-CoV-2 Antigen assay.

|  | cobas SARS-CoV-2 assay | N total | Elecsys SARS-CoV-2<br>Antigen negative | Elecsys SARS-CoV-2<br>Antigen positive | Relative sensitivity/specificity, %<br>(95%, CI two-sided) |
| --- | --- | --- | --- | --- | --- |
| **Relative**<br>**sensitivity** | **PCR positive** | **392** | **156** | **236** | **60.2**<br>**(55.2–65.1)** |
|  | >10<sup>4</sup> copies/mL (~Ct <29.9) | 223 | 14 | 209 | 93.7<br>(89.7–96.5) |
|  | >10<sup>5</sup> copies/mL (~Ct <26.6) | 171 | 0 | 171 | 100<br>(97.9–100) |
|  | >10<sup>6</sup> copies/mL (~Ct <23.0) | 122 | 0 | 122 | 100<br>(97.0–100) |
| **Relative**<br>**specificity** | **PCR negative** | **2747<sup>a</sup>** | **2743** | **4** | **99.9**<br>**(99.6–100)** |

<sup>a</sup>15 samples invalid with cobas SARS-CoV-2 RT-PCR, but negative with another SARS-CoV-2 RT-PCR test.
CI, confidence interval; Ct, cycle threshold; N, number of samples

For samples from symptomatic patients, the assay reached an overall relative sensitivity of 68.4% (95% CI 62.9–73.5, N=313), increasing to 83.2% (95% CI 76.2–88.8, N=149) if symptoms had manifested within the last 5 days (**Table S2**). On the other hand, the overall sensitivity observed in asymptomatic patients was only 27.8% (95% CI 18.3–39.1, N=79). Relative sensitivities corresponded with high RNA levels and gradually decreased as RNA levels decreased. A considerable increase in sensitivity was observed when analysis was limited to samples containing >10,000 copies/mL, reaching 95.9% (95% CI 92.2–98.2, N=197) for symptomatic patients and 76.9% (95% CI 56.4–91.0, N=26) for asymptomatic patients (**Table S2**).

To analyze the overall assay performance, samples were sorted by SARS-CoV-2 RNA copies/mL and individual sensitivities calculated for sets of 20 samples (shown using a heat map in **Fig. 2A**). Of note, the sensitivity of the assay was 95% and 50% at concentrations of around 2×10^5^ and 2×10^4^–5×10^3^ viral copies/mL, respectively, but <10% for samples with <10^3^ copies/mL. When the analysis was focused on samples containing RNA levels beyond 95% sensitivity of the immunoassay (10,000 copies/mL; Ct <29.9), a significant linear correlation was observed between the Elecsys SARS-CoV-2 Antigen assay COI value and SARS-CoV-2 RNA copies/mL (p=<0.0001; r^2^=0.786; **Fig. 2B**). Also, in this group, a sensitivity of 98.3% (95% CI 94.0–99.8) was achieved in the sub-analysis of patients tested less than 5 days after symptom onset.

**Figure 2.**
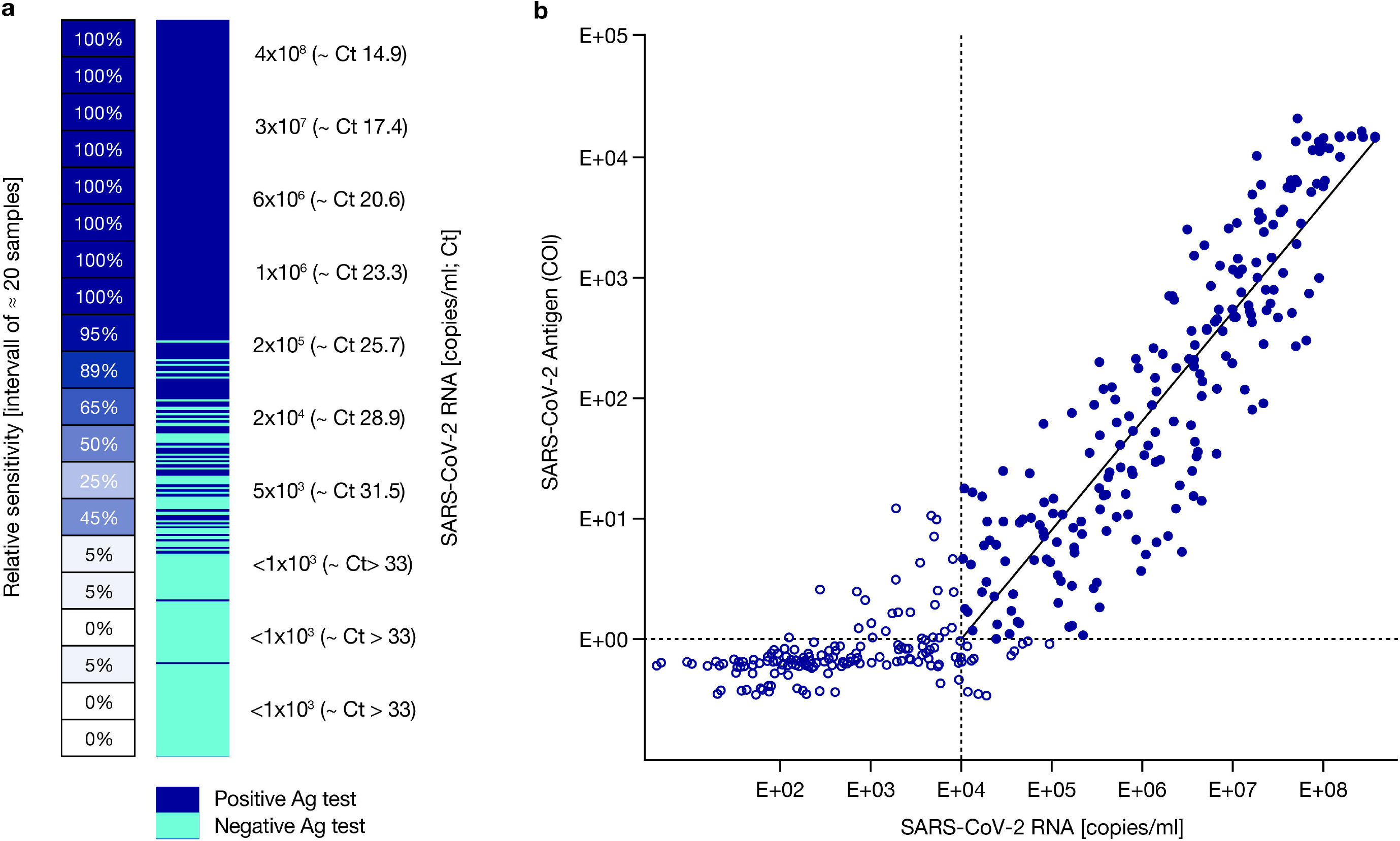
**a)** Elecsys SARS-CoV-2 Antigen assay results in relation to cobas SARS-CoV-2 RNA copies/mL. Dark blue fields represent positive and light blue fields represent negative Elecsys SARS-CoV-2 Antigen test results. On left side, sensitivity was calculated every ≈ 20 samples included in the interval of the heatmap (copies/mL); **b)** Correlation between Elecsys SARS-CoV-2 Antigen assay (log10 [COI], y-axis) and cobas SARS-CoV-2 PCR assay (log10 [RNA copies/mL], x-axis). Closed blue circles: measurements within linear range considered for the trendline and correlation (SARS-CoV-2 RNA ≥10^5^ copies/mL; COI ≥1). Open blue circles: measurements outside of the linear range of Elecsys SARS-CoV-2 or one cobas SARS-CoV-2 PCR Target negative, not considered for trendline and correlation. p=<0.0001; r^2^ =0.786. Ag, antigen; COI, cut-off index; Ct, cycle threshold

## DISCUSSION

The introduction of rapid SARS-CoV-2 antigen testing into the market was met with a lot of initial enthusiasm due to the promise of broad availability and easy application. While there are many different methods for antigen detection in clinical practice, such as enzyme-linked immunosorbent assays and ECLIAs, the first wave of commercial tests consisted predominantly of rapid, point-of-care LFTs with optical read-outs. Preliminary performance data demonstrated good sensitivity in high-viral-load samples, but low concentrations of virus are frequently missed, thus leading to the sobering conclusion that antigen tests cannot provide the same definitive results as RT-qPCR [5, 11-13]. Although PCR tests are analytically superior, their effectiveness for infection control has often been hampered by supply shortages and reporting delays.

Recent studies have shown a correlation between the probability of recovering infectious virus from clinical samples and the time of onset of symptoms, as well as RNA concentrations (>1 ×10^6^ copies/mL) within the specimen [14, 15]. This suggests that the risk of transmission is highest during the early stages of disease, when viral titers are at their peak, which would also facilitate detection by antigen test. A recent study by McKay et al. [16] demonstrated good correlation between positive antigen-test and positive viral culture; however, negative viral culture should not be misinterpreted as a reliable indicator for ruling out transmission risk. It has further been suggested that a low sensitivity test can be just as effective at detecting infections as a highly sensitive one, if testing can be made possible at increased frequency [6]. Most currently available SARS-CoV-2 antigen tests utilize a rapid, point-of-care LFT format, thus relying on a high degree of human interaction. They are unsuitable for automation and additional human operators are required at the same proportion as testing capacity is scaled up. While there are clear benefits of a point-of-care test that can be carried out with minimal training and requiring no specialized equipment [17], testing en masse (e.g. in healthcare facilities) is vastly more efficient when performed automatically using high-throughput immunoassay platforms.

In this study, we stratified samples to assess the relative sensitivity of the new Elecsys SARS-CoV-2 Antigen assay for the cobas immunoassay analyzer systems using the gold standard, RT-qPCR. Although the overall relative sensitivity for the entire sample set was 60.2%, among those patients that were tested in the first 5 days from the onset of symptoms, the sensitivity increased to 83.2%. Furthermore, the Elecsys SARS-CoV-2 Antigen assay was able to detect 93.7% of patients with elevated risk of transmission (Ct <30 and accordingly c(RNA)> 10,000 copies/mL). Overall relative specificity was determined as 99.9% in a total of 2747 negative samples. These results are in line with existing preliminary data on the clinical performance of other available SARS-CoV-2 antigen tests [12, 18], including other high-throughput SARS-CoV-2 antigen tests like the Lumipulse [19], demonstrating a cumulative sensitivity of 55.2% compared with PCR.

The main limitation of the present study is that it is not a head-to-head comparison of the different systems on the market, and future investigation is needed to address this concern. However, the high-throughput Elecsys SARS-CoV-2 Antigen assay might be a valuable analytic option in the coming months in the struggle to control the pandemic.

## CONCLUSIONS

The novel Elecsys SARS-CoV-2 Antigen immunoassay showed good performance in a broad set of clinical samples compared to RT-PCR. It demonstrated a high reliability in detecting samples containing high concentrations of viral RNA, indicative of high transmission potential. Furthermore, a relative specificity of 99.9% ensures a minimal rate of false-positives, consequently saving resources on confirmation-testing and unnecessary quarantine. The ability to run the test on high-throughput immunoassay platforms, and a time-to-result of approximately 18 minutes, allow for en masse deployment of SARS-CoV-2 antigen testing, for example in the context of centralized large-scale testing schemes.

## Funding

This study was funded by Roche Diagnostics GmbH (Mannheim, Germany).

## Authorship

All named authors meet the International Committee of Medical Journal Editors (ICMJE) criteria for authorship for this article, take responsibility for the integrity of the work as a whole, and have given their approval for this version to be published.

## Authorship Contributions

Tina Laengin, Tanja Schneider, and Kathrin Schönfeld developed the study conceptualization and design and study protocol, and managed the study conduct, database generation and data validation. Marc Lütgehetmann, Stojan Perisic, and Matthias F. Bauer provided sample material and Marc Lütgehetmann, Stojan Perisic, and Elena Riester generated data during study testing. Marc Lütgehetmann, Tanja Schneider, and Tina Laengin conceived the original idea of the manuscript. Dominik Nörz, Flaminia Olearo, Marc Lütgehetmann, and Tanja Schneider formally analyzed the data and its representations for the manuscript. Dominik Nörz, Marc Lütgehetmann, and Flaminia Olearo drafted the manuscript and interpreted the data with critical input from Tanja Schneider and Tina Laengin. All authors reviewed and revised the manuscript and approved it for publication.

## Medical Writing, Editorial, and Other Assistance

The authors thank: Serology team of the University Medical Center Hamburg-Eppendorf, Institute of Medical Microbiology, Virology and Hygiene; the laboratory team of the Institute of Laboratory Diagnostics, Hygiene and Transfusion Medicine at the Klinikum Ludwigshafen; the Corona specimen acquisition team and its Coordinator Mr. OA Christian Menzel (Clinic of Stuttgart), Corona testing facility team (Clinic of Stuttgart) as well as Ms. Pauline Weissleder and Ms. Sina Semenowitsch (Clinic of Stuttgart) for great help with specimen handling and data processing; Sigrid Reichhuber, Janina Edion, and Yvonne Knack (Roche Diagnostics) for management of investigation sites, data acquisition, and study monitoring. Medical writing support, under the direction of the authors, was provided by Tina Patrick (Elements Communications Ltd, Westerham, Kent, UK) and was funded by Roche Diagnostics International Ltd, Rotkreuz, Switzerland.

COBAS, COBAS E and ELECSYS are trademarks of Roche.

## Disclosures

Tanja Schneider, Kathrin Schönfeld, and Tina Laengin are employees of Roche Diagnostics GmbH. Marc Lütgehetmann has received speaker’s honoraria and related travel expenses from Roche Diagnostics. Elena Riester has received speaker’s honorarium from Roche. Stojan Perisic, Flaminia Olearo, Matthias F. Bauer and Dominik Nörz have no conflicts to report.

## Compliance with Ethics Guidelines

This study was conducted in accordance with applicable regulations, the study protocol provided by Roche Diagnostics, and the principles of the Declaration of Helsinki of 1964, and its later amendments. The use of anonymized remnant samples material was approved by ethical review committees prior to study initiation (ethics committee names and approval numbers: Ethik-Kommission bei der Landesärztekammer Baden-Württemberg; F-2020-154 [Stuttgart]; Ethik-Kommission der Ärztekammer Hamburg: WF-184/20 [Hamburg]; Ethik-Kommission bei der Landesärztekammer Rheinland-Pfalz: 2020-15449[Ludwigshafen]).

## Data Availability

The datasets generated during and/or analyzed during the current study are available from the corresponding author on reasonable request.

## Notes

### Competing Interest Statement

Tanja Schneider, Kathrin Schoenfeld, and Tina Laengin are employees of Roche Diagnostics GmbH. Marc Luetgehetmann has received speaker's honoraria and related travel expenses from Roche Diagnostics. Elena Riester has received speaker's honorarium from Roche. Stojan Perisic, Flaminia Olearo, Matthias F. Bauer and Dominik Noerz have no conflicts to report. This study was funded by Roche Diagnostics GmbH (Mannheim, Germany). Medical writing support, under the direction of the authors, was provided by Tina Patrick (Elements Communications Ltd, Westerham, Kent, UK) and was funded by Roche Diagnostics International Ltd, Rotkreuz, Switzerland.

### Funding Statement

This study was funded by Roche Diagnostics GmbH (Mannheim, Germany). Medical writing support, under the direction of the authors, was provided by Tina Patrick (Elements Communications Ltd, Westerham, Kent, UK) and was funded by Roche Diagnostics International Ltd, Rotkreuz, Switzerland.

### Author Declarations

This study was conducted in accordance with applicable regulations, the study protocol provided by Roche Diagnostics, and the principles of the Declaration of Helsinki of 1964, and its later amendments. The use of anonymized remnant samples material was approved by ethical review committees prior to study initiation (ethics committee names and approval numbers: Ethik-Kommission bei der Landesarztekammer Baden-Wurttemberg; F-2020-154 [Stuttgart]; Ethik-Kommission der Arztekammer Hamburg: WF-184/20 [Hamburg]; Ethik-Kommission bei der Landesarztekammer Rheinland-Pfalz: 2020-15449[Ludwigshafen]).

