## Supplemental Table 1 and Table 2 for "Multicenter evaluation of a fully automated high-throughput SARS-CoV-2 antigen immunoassay"

**Supplementary Material**

**Table S1**. List of origin of samples, sites, and devices used for testing.

| **Sample provision** | **Samples, N total (N PCR-positive)** |
| --- | --- |
| Stuttgart | 1331 (95) |
| Hamburg | 1058 (58) |
| Ludwigshafen | 647 (151) |
| Commercial vendor | 103 (88) |
| **Testing site** | **Samples, N total** |
| Hamburg (cobas e 411) | 1808 |
| Augsburg (cobas e 801) | 1331 |

Ct, cycle threshold; N, number of samples

**Table S2.** Analytic performance of Elecsys SARS-CoV-2 Antigen stratified by symptoms, duration of symptoms, and according to viral load (copies/mL).

|  | **Description** | **N total** | **Elecsys SARS-CoV-2 Antigen negative** | **Elecsys SARS-CoV-2  Antigen positive** | **Relative sensitivity, % (95%, CI two-sided)** |
| --- | --- | --- | --- | --- | --- |
| **Symptomatic** | **All** | **313** | **99** | **214** | **68.4 (62.9–73.5)** |
|  | *Symptomatic <5 days DPSO*  *Symptomatic >5 DPSO*  *Symptomatic with unknown DPSO* | *149*  *109*  *55* | *25*  *59*  *15* | *124*  *50*  *40* | *83.2 (76.2–88.8)*  *45.9 (36.3–55.7)*  *72.7 (59.0–83.9)* |
|  | **T2 >10^4^ copies/mL (~Ct 29.9)** | **197** | **8** | **189** | **95.9 (92.2–98.2)** |
|  | *Symptomatic <5 days DPSO*  *Symptomatic >5 DPSO*  *Symptomatic with unknown DPSO* | *118*  *42*  *37* | *2*  *5*  *1* | *116*  *37*  *36* | *98.3 (94.0–99.8)*  *88.1 (74.4–96.0)*  *97.3 (85.8–99.9)* |
|  | **T2 >10^5^ copies/mL (~Ct 26.6)** | **153** | **0** | **153** | **100 (97.6–100)** |
|  | *Symptomatic <5 days DPSO*  *Symptomatic >5 DPSO*  *Symptomatic with unknown DPSO* | *99*  *28*  *26* | *0*  *0*  *0* | *99*  *28*  *26* | *100 (96.3–100)*  *100 (87.7–100)*  *100 (86.8–100)* |
|  | **T2 >10^6^ copies/mL (~Ct 23.0)** | **109** | **0** | **109** | **100 (96.7–100)** |
|  | *Symptomatic <5 days DPSO*  *Symptomatic >5 DPSO*  *Symptomatic with unknown DPSO* | *75*  *20*  *14* | *0*  *0*  *0* | *75*  *20*  *14* | *100 (95.2–100)*  *100 (83.2–100)*  *100 (76.8–100)* |
| **Asymptomatic**  **(Known / suspected exposure + screening)** | **All** | 79 | 57 | 22 | 27.8 (18.3*–*39.1) |
|  | **T2 >10^4^ copies/mL (~Ct 29.9)** | 26 | 6 | 20 | 76.9 (56.4*–*91.0) |
|  | **T2 >10^5^ copies/mL (~Ct 26.6)** | 18 | 0 | 18 | 100 (81.5*–*100) |
|  | **T2 >10^6^ copies/mL (~Ct 23.0)** | 13 | 0 | 13 | 100 (75.3*–*100) |

CI, confidence interval; Ct, cycle threshold; DPSO, days post symptom onset; N, number of samples; T2, Target-2
